# Seeing the Aging Heart: Multimodal AI Quantifies Cardiac Biological Aging from Angiography, Echocardiography, and ECG

**DOI:** 10.1101/2025.10.27.25338901

**Authors:** Jeffrey G. Malins, Behrouz Rostami, Eunjung Lee, Puskar Bhattarai, DM Anissuzaman, John I. Jackson, Seda Camalan, Timothy J. Poterucha, Kenneth A. Fetterly, Mohamad Alkhouli, Amir Lerman, Suraj Kapa, Matthew R. Callstrom, Garvan C. Kane, Francisco Lopez-Jimenez, Paul A. Friedman, Zachi I. Attia

## Abstract

Cardiac biological aging results in vascular, structural, and electrical changes that account for age-related cardiovascular disease. Using techniques such as deep neural networks, Artificial Intelligence (AI) based analysis of signals such as ECGs and echocardiograms has revealed that patients age at different rates, and that differences between AI-estimated age and chronological age (“age gaps”) are associated with long-term survival. However, so far, cardiac biological aging has focused on single modality estimation and in addition has not yet been evaluated using coronary angiograms. In this study, we first developed a cardiac age estimation model using coronary angiograms. Then, in a group of 1,345 patients who had an echocardiogram and ECG within one month of their angiogram, we examined how patient survival was associated with age gaps in each modality in isolation (using previously developed models for echocardiograms and ECGs) and then across the three modalities combined. For the average across the three modalities, we observed a hazard ratio (HR) of 2.24 (95% confidence interval: 1.71-2.92) per unit increase in the age gap, which was a marked increase compared to each modality on its own (HR of 1.63, 1.54, and 1.24 for angiograms, echocardiograms, and ECGs respectively). This result demonstrates that the predictive value of AI-estimated cardiac age compounds with additional inputs. While angiograms are not practical for routine monitoring, they serve as proof of concept that richer vascular imaging can enhance biological age prediction. As interventions targeting aging emerge, we will need objective tools to measure their impact. Multi-modal cardiac age may provide a scalable, interpretable marker of cardiovascular aging and possibly even rejuvenation.

## Introduction

Chronological age is an imprecise surrogate for physiological aging. While it is widely used in clinical decision-making and risk scoring, it does not capture the significant interindividual and organ-specific variability associated with aging. Cardiovascular disease reflects this disconnect: patients of the same chronological age often differ widely in cardiac structure, function, and outcomes. Artificial Intelligence (AI) offers an alternative way of conceptualizing age. For example, using either electrocardiogram (ECG) or echocardiographic data, patient age can be reliably estimated using deep learning models [1–4]. Using these estimates, ‘aging clock’ models can be applied to examine the extent to which the discrepancy between chronological age and model-estimated age acts as a biomarker to predict mortality and acts as a potential therapeutic metric for age-related diseases [5].

Although AI models have been developed to estimate measurements such as left ventricular ejection fraction from routine coronary angiograms [6, 7], deep learning has not yet been applied to these images to estimate patient age. This modality, however, offers rich vascular information that is distinct from electrical (ECG) and structural (echocardiographic) signals and may provide additional insight into cardiac aging [8]. Therefore, in the current study, our first aim was to evaluate the extent to which deep learning models can estimate patient age from routine coronary angiographic images.

Once we developed the new angiogram-based AI-age estimation model, our second aim was to then compare AI-age estimates from this model with AI-age estimates obtained for the same patients using echocardiogram- and ECG-based AI-age deep learning models previously developed by our group [1, 9]. Based on previous work that has used multiple cardiac imaging modalities to triangulate sources of cardiac aging [10, 11], we hypothesized that each modality captures different aspects of cardiac aging, and therefore age estimates would be only weakly related across modalities. Our final aim was to then evaluate whether considering the three modalities in tandem would better account for patient survival outcomes than considering each modality in isolation. We hypothesized that combining multimodal data streams to assess vascular (angiography), structural (echocardiography), and electrical (ECG) changes would better account for patients’ long-term survival.

## Materials and Methods

### Angiogram-Based Algorithms

#### Dataset

All study procedures were approved by the Mayo Clinic Institutional Review Board, and patient cohorts only included individuals who had provided prior authorization for the use of their data in research. The dataset used in this study was a set of angiogram cine images collected between 2015 to 2021 at the Minnesota, Arizona, and Florida sites of Mayo Clinic. Across and within these sites, the images are different in some aspects such as their dimensions because they were captured by different imaging instruments associated with different manufacturers.

Starting from a collection of 813,500 angiogram clips collected from 28,075 patients, invalid cases (invalid number of frames and missing ID) as well as cases that were not authorized to be used in research were excluded, and only studies with a cine rate of 15 frames/sec were kept. This filtering process resulted in a final cohort of 208,918 angiogram clips from 26,758 distinct patients (31,535 studies, as some patients had more than one study date included in the dataset). Ultimately, the cohort was split randomly into training, validation, and testing sets, with respective proportions of 70%, 15%, and 15% of the main cohort. This was done at the patient level, meaning that no patient appeared in more than one subset.

Note that for the dataset to be as comprehensive as possible, patients with a heart transplant were not excluded. Nor were patients excluded if they had a prosthesis, coronary artery bypass grafting (CABG), or prior stents/other devices.

#### Data preparation

From each DICOM file, 16 consecutive frames starting from the 16^th^ frame were extracted and stored in .png format to be used for model training and evaluation. The first 15 frames of each video clip were discarded in an attempt to remove images corresponding to the time of filling with an iodine-based contrast agent. During training and before presenting video clips to the model, multiple augmentation transformations were applied to the clips, including resizing, random rotation, and horizontal and vertical flipping.

#### Model training

Routine coronary angiogram images/videos were utilized for model training. More specifically, three different approaches were used: single image input, multi-frame input, and video clip input (Figure 1). In the single image input strategy, the input to the model was a single 2D image (frame) that was selected from the center of the video clip. A 2D version of the ResNet architecture [12] was used for the model, and the output for each data sample was obtained by applying the model on the selected frame. For the multi-frame input strategy, a new 2D ResNet model (with 152 layers) was trained, with the input consisting of four frames that were selected at regular intervals from each video clip in our dataset. The model was applied on each frame, and the overall outcome of the model was obtained by taking the average of these four outputs. For the video-based strategy, a 3D ResNet architecture (with 152 layers) was instead used [12], and the input to the model for each data sample was a 16-frame video clip. In all cases, the ground truth age used for training was a decimal value reflecting the difference between the patient’s date of birth and their angiogram visit date.

**Figure 1.**
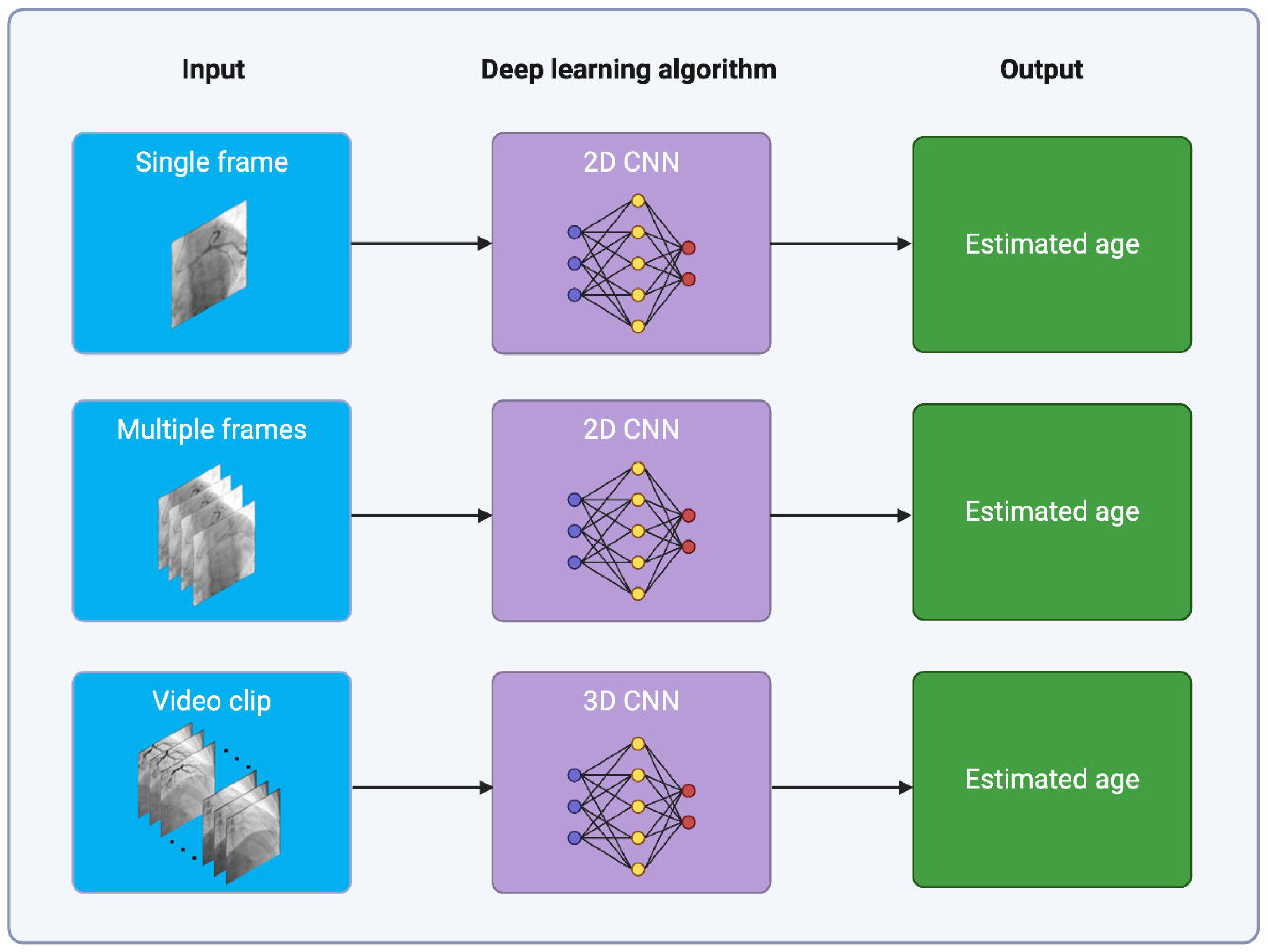
The three input strategies for the angiogram-based models evaluated in the current study. CNN = convolutional neural network

#### Model hyperparameters

Each model was trained for 50 epochs, and the value of 1e-5 was used for the initial learning rate, which was then decayed during training using the ReduceLROnPlateau strategy [13]. A value of 0.001 was selected for weight decay, and the mini-batch size was set to 64. The Adam algorithm [14] was utilized for optimization, and the loss functions was Root Mean Square Error (RMSE). All experiments and models were implemented in the Python programming language (version 3.10.4) and on the PyTorch (version 1.12.1+cu116) platform [15] using two NVIDIA GA102GL [A10] GPUs.

#### Model evaluation

Model performance was evaluated by applying trained models on the testing set. Root Mean Squared Error (RMSE) along with R-squared (coefficient of determination) were utilized as model evaluation metrics. More details about these metrics can be found in [16].

### Comparison of age estimates across cardiac modalities

From the testing dataset of the angiogram-based age estimation model, a subset of 1,345 distinct patients was selected by restricting the set to only those who had a transthoracic echocardiogram as well as least one electrocardiogram within 30 days of their angiogram. If a patient had more than one echocardiographic exam, the exam closest to the angiogram date was chosen, whereas if they had more than one ECG within the 30 days, all ECGs were chosen. Age estimates were then computed separately for each modality as follows: for angiograms, the multi-frame 2D CNN approach was used, and average estimates across frames were computed; for echocardiograms, estimates were obtained using a previously developed view-agnostic modeling approach for videos [9], which takes an average estimate across all views containing the left ventricle (as determined using a view classifier; [17]); for electrocardiograms, estimates were obtained for 12-lead signals using AI models previously developed by our team [1], with average estimates taken across ECGs if there was more than one ECG within 30 days of the angiogram. To avoid data leakage, patients who were part of the training or validation datasets for the AI-echo, AI-ECG, or echocardiographic view classifier models were excluded prior to arriving at the final set of 1,345 patients. For this set of patients, pairwise comparisons were then performed to compare model estimates across modalities. When performing these comparisons, respective age gaps for each of the modalities were computed by taking the difference between a patient’s birth date and the date of each specific visit.

### Evaluating the relationship between AI-age estimates and patient survival

First, we examined patient survival curves for patients from the testing set of the angiogram-based AI-age model from whom survival outcomes were available (*N* = 4,009 patients out of 4,015 in the testing set). Then, we examined patient survival curves for the subset of 1,345 patients with all three tests: angiography, echocardiography, and ECG. Note that for this analysis, for patients with multiple ECGs within the time window, chronological age was defined at the date of the most recent ECG, which was used as the survival analysis baseline.

When performing these analyses, for the purposes of visualization, we grouped patients according to whether the discrepancy between chronological age and model-estimated age was within 1 SD of the mean discrepancy (“within 1 SD”), or whether the model-estimated age was greater than 1 SD above the mean discrepancy (“1+ SD older”) or greater than 1 SD below the mean discrepancy (“1+ SD younger”). Note that means and SDs were computed separately for each respective modality. We then performed Cox proportional hazards regression models in R version 4.4.1, adjusting for chronological age and patient sex (*survival* library 3.7-0 [18], visualized using *survminer* 0.4.9 [19] and *adjustedCurves* 0.11.1 [20]), treating the “within 1 SD” group as the reference group. For the combined model across modalities, we grouped patients by tallying the number of modalities out of the three for which a patient was estimated to be at least 1 SD older, within 1 SD of their chronological age, or 1 SD younger (i.e., considered older by at least two modalities; 2 within, 1 older; all 3 within; and so on).

For statistical analysis, we instead took the continuous value of the discrepancy between chronological age and model-estimated age (termed as the ‘age gap’ throughout) and normalized this value within each modality, by first subtracting the mean of the age gap and then by dividing by the standard deviation of the age gap. This normalized age gap was then used as a regressor in Cox proportional hazards regression models. For the combined model, we instead took the average normalized age gap across modalities and used this as a regressor. All models adjusted for chronological age and patient sex (with female as the reference level for this factor).

## Results

### Sample characteristics

Table 1 provides demographic and clinical characteristics of the patient cohorts for training (*N* = 18,730), validation (*N* = 4,013), and testing (*N* = 4,015) of the angiogram-based model for age estimation, as well as the subset of patients who had an echocardiogram and at least one ECG within thirty days of their angiogram (*N* = 1,345). In general, across the cohorts, mean age was 67, 33-35% of patients were female, and there was a high prevalence of cardiac pathologies and related comorbidities.

**Table 1.** Demographic and clinical characteristics of the patient cohorts used for model development and testing.

|  | Angiogram Model Development |  |  | Subset of testing dataset with echo and ECG within 30 days of angiogram |
| --- | --- | --- | --- | --- |
|  | Training | Validation | Testing |  |
| Number of studies | 22,046 | 4,722 | 4,767 | 1,345 |
| Number of unique patients | 18,730 | 4,013 | 4,015 | 1,345 |
| Age on study date (years) |  |  |  |  |
| mean ± standard deviation | 67 ± 12 | 67 ± 12 | 67 ± 13 | 67 ± 13 |
| range | 18-102 | 18-98 | 18-99 | 19-97 |
| Sex (%) |  |  |  |  |
| female | 34.81 | 33.86 | 34.74 | 35.54 |
| male | 65.19 | 66.14 | 65.26 | 64.46 |
| Race and ethnicity (%) |  |  |  |  |
| NH American Indian / Alaska Native | 0.39 | 0.37 | 0.45 | 0.45 |
| NH Asian | 1.38 | 1.35 | 1.44 | 1.04 |
| NH Black / African American | 2.35 | 2.92 | 2.79 | 1.56 |
| Hispanic / Latino | 2.53 | 2.59 | 3.21 | 2.01 |
| NH Native Hawaiian / Other Pacific Islander | 0.10 | 0.02 | 0.17 | 0.30 |
| NH White | 89.80 | 89.14 | 88.59 | 91.38 |
| Other or unknown | 3.45 | 3.61 | 3.34 | 3.27 |
| Comorbidities (% positive) |  |  |  |  |
| Hypertension | 74.58 | 74.73 | 75.02 | 72.49 |
| Atrial fibrillation | 30.81 | 30.77 | 30.41 | 28.62 |
| Hyperlipidaemia | 72.40 | 71.67 | 72.60 | 71.23 |
| Congestive heart failure | 44.85 | 43.76 | 46.75 | 44.54 |
| Myocardial infarction | 34.95 | 34.56 | 34.07 | 33.53 |
| Valvular disease | 57.92 | 56.62 | 57.14 | 52.57 |
| Peripheral vascular disorders | 44.97 | 43.28 | 45.13 | 40.30 |
| Chronic pulmonary disease | 34.90 | 33.94 | 35.79 | 33.68 |
| Diabetes mellitus | 33.30 | 34.81 | 34.57 | 30.78 |
| Renal failure | 28.62 | 29.23 | 29.34 | 26.62 |
| Liver disease | 16.30 | 16.80 | 17.06 | 13.61 |

*Note.* Binary sex as well as race and ethnicity were self-reported and obtained from each patient's medical record. Comorbidities were ascertained using the *comorbidity* package in R [20][20] based on ICD-9 or ICD-10 codes noted in the patient's medical record up to and including the angiogram study date. For patients in the angiogram model development datasets, which contained numerous patients with more than one study date, the earliest study was taken when reporting age and comorbidities. NH = non-Hispanic; echo = echocardiogram

### Age estimation using angiogram-based models

Figure 2 displays scatterplots as well as Bland-Altman plots for the age estimation experiments. The best outcome was observed for the multi-frame approach, which had a root mean squared error (RMSE) of 8.317 and a coefficient of determination value of 0.582.

**Figure 2.**
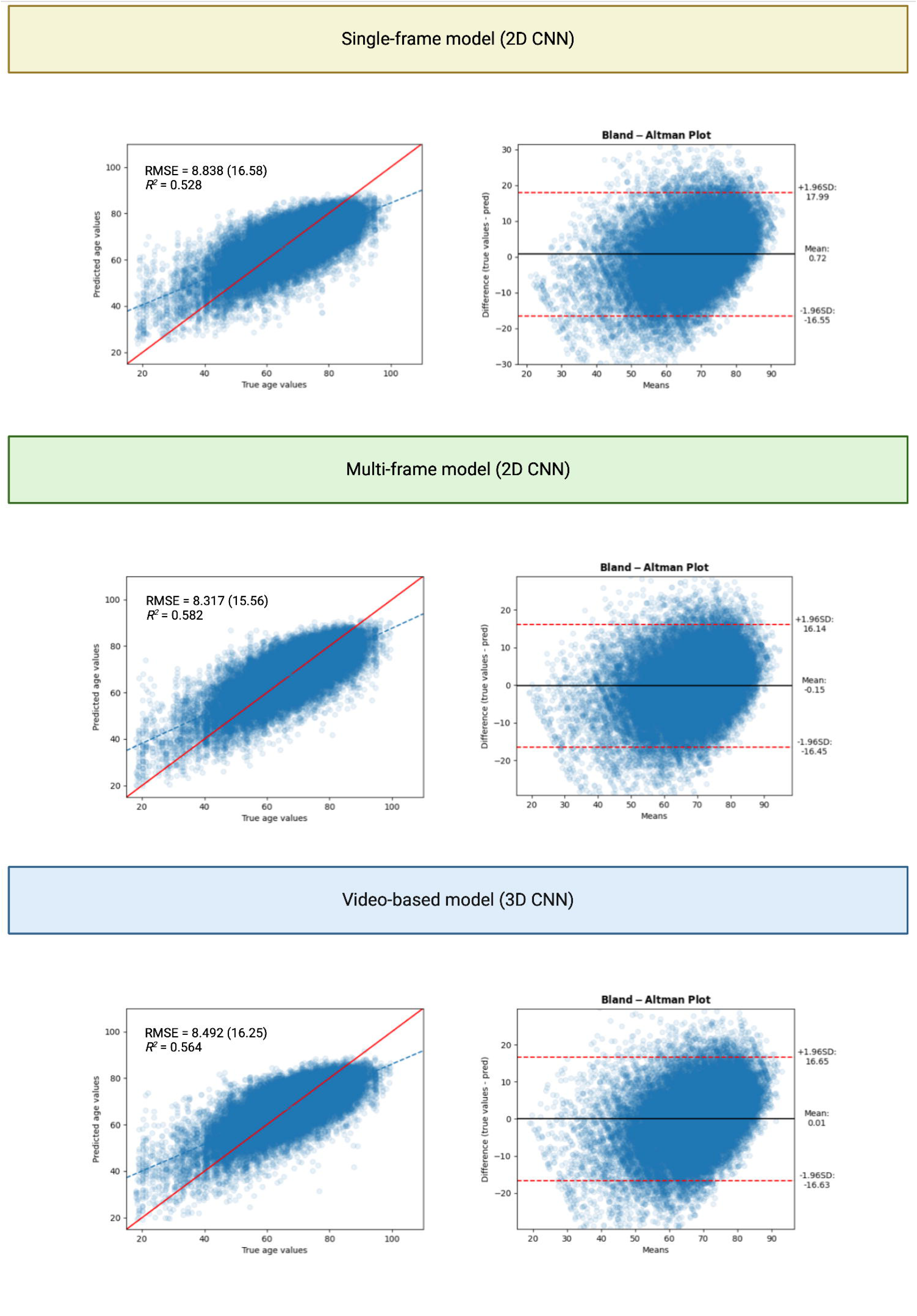
Scatter and Bland-Altman plots for the age estimation experiments for the angiogram-based models. CNN = convolutional neural network; RMSE = root mean squared error

As shown in Figure 2, there is a systematic bias toward the mean, such that patient age tends to be overestimated in younger patients and underestimated in older patients. This is reflected in the slope of the regression line – which is shallower than that of the identity line – as well as the Bland-Altman plots. With that said, in Figure 3, which displays the ROC curves obtained when different age cutoffs were applied to threshold continuous age estimates into binary outputs (i.e., below or equal to 40 versus over 40, and so on), the age estimation model still performs well at all age levels, albeit slightly poorer for older patients.

**Figure 3.**
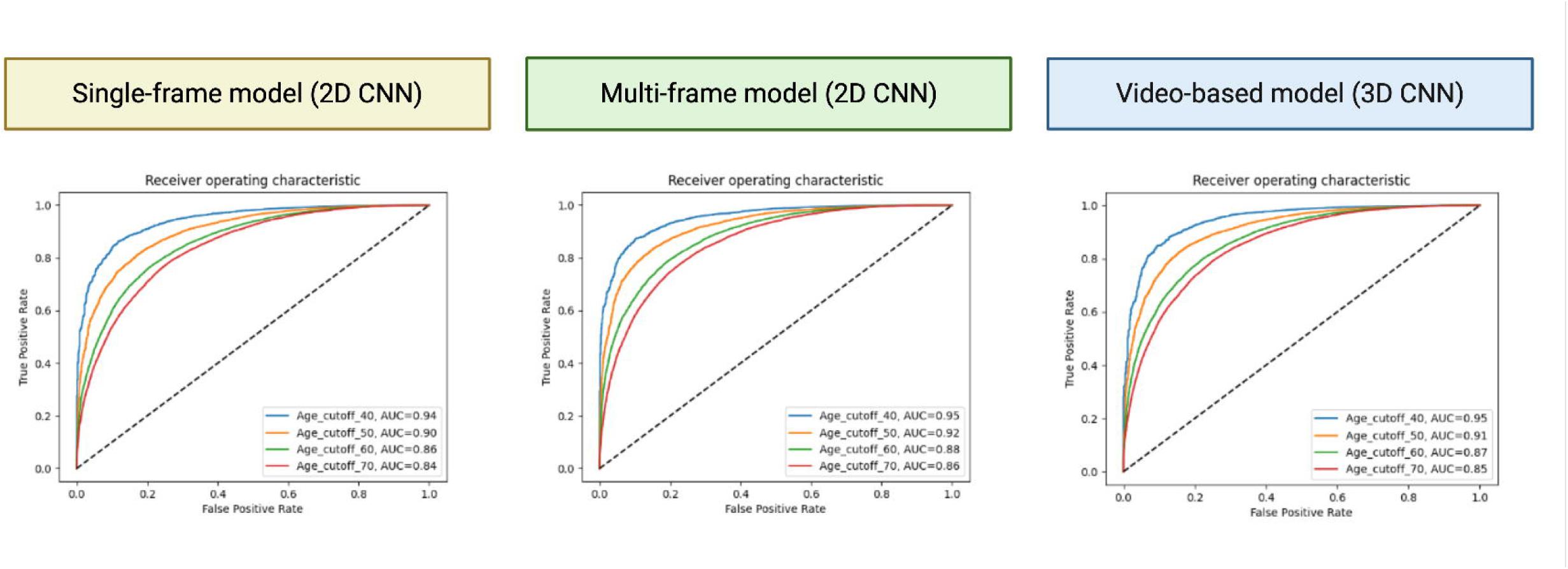
ROC curves for age classification using estimated age values from the angiogram-based models. CNN = convolutional neural network; AUC = area under the curve

### Comparison of age estimates across cardiac modalities

Figure 4 shows the results from pairwise correlations across modalities for the latency difference between chronological age and model-estimated age (*N* = 1,345 patients). As shown in the figure, age estimation performance was comparable between the AI-angiogram (*r* = 0.862) and AI-echo (*r* = 0.876) models yet inferior for the AI-ECG model (*r* = 0.676). Differences between modalities were also apparent in the strength of the pairwise correlations between age gaps, which was strongest for angiography and echocardiography (*r* = 0.636), intermediate for echocardiography and ECG (*r* = 0.619), and weakest for angiography and ECG (*r* = 0.570).

**Figure 4.**
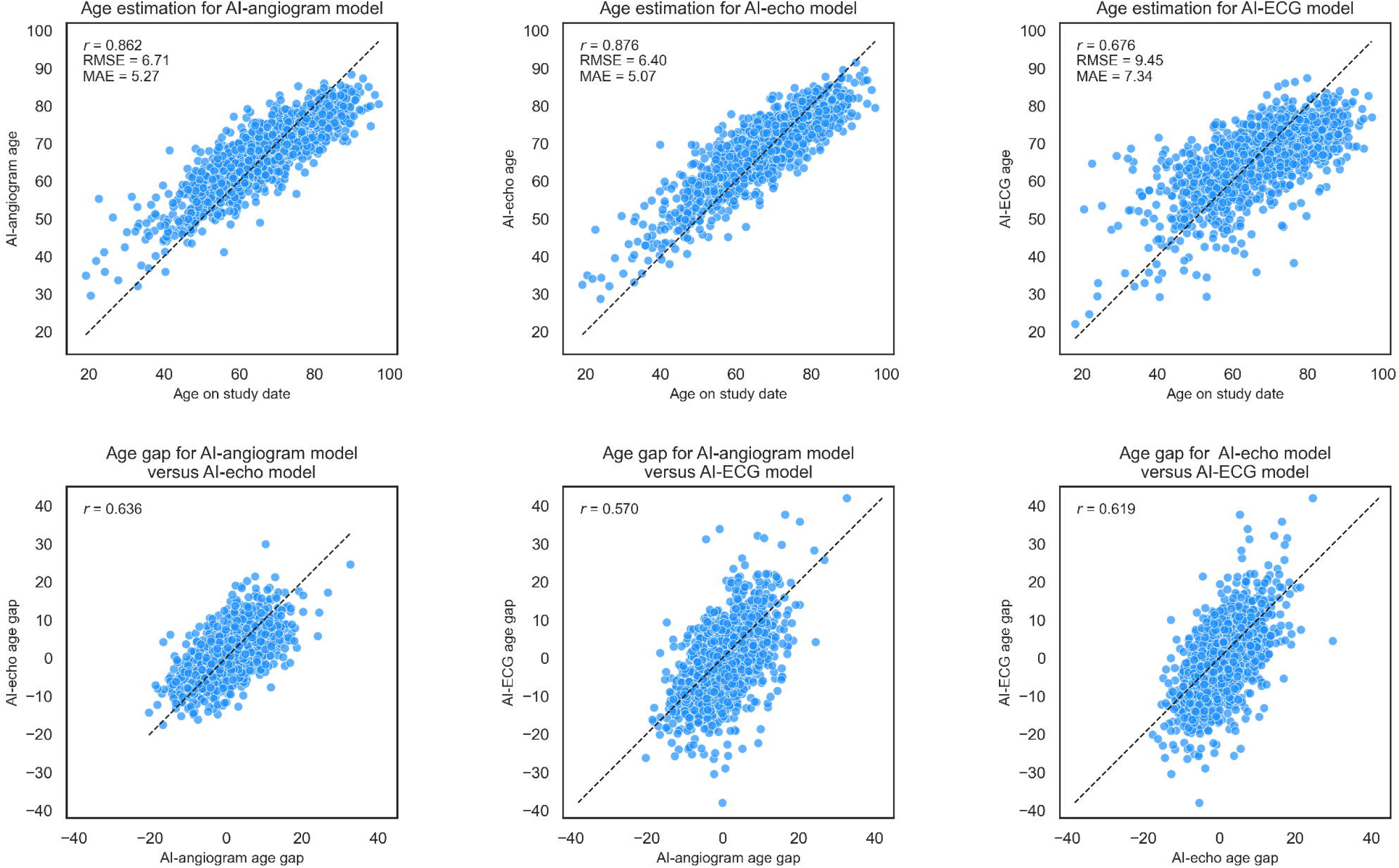
*Top row*: model-estimated age compared to chronological age for the AI-angiogram, AI-echo, and AI-ECG models respectively. *Bottom row*: pairwise correlations for age gaps (i.e., differences between chronological age and model-estimated age) across modalities. echo = echocardiogram

These pairwise correlations across modalities were notably weaker in strength compared to the correlations of the per-modality estimates themselves with ground truth age. The lack of correlation in the age gap data suggests that each modality contributes novel information to the age estimates.

### Evaluating the relationship between AI-age estimates and patient survival

Survival curves for individual modalities are shown in Figure 5 for both the full testing dataset from the angiogram-based model (*N* = 4,009) as well as the subset of patients from whom both echocardiograms and ECGs were recorded within 30 days of the angiogram study date (*N* = 1,345). As shown in the figure, for the angiogram-based model, the hazard ratio (HR) for the normalized age gap was 1.43 (95% CI of 1.29-1.58, *p* < 0.0001) for the full testing dataset, meaning that as the average normalized age gap increased by one unit (i.e., the average of the models estimating a patient’s age to be higher than their chronological age), there was a 43% increased hazard of death. For the subset of patients with all three modalities, the HR for the normalized age gap was 1.63 (1.35-1.97, *p* < 0.0001) for the angiogram-based model, 1.54 (1.30-1.83, *p* < 0.0001) for the echocardiogram-based model, and 1.24 (1.02-1.50, *p* = 0.029) for the ECG-based model.

**Figure 5.**
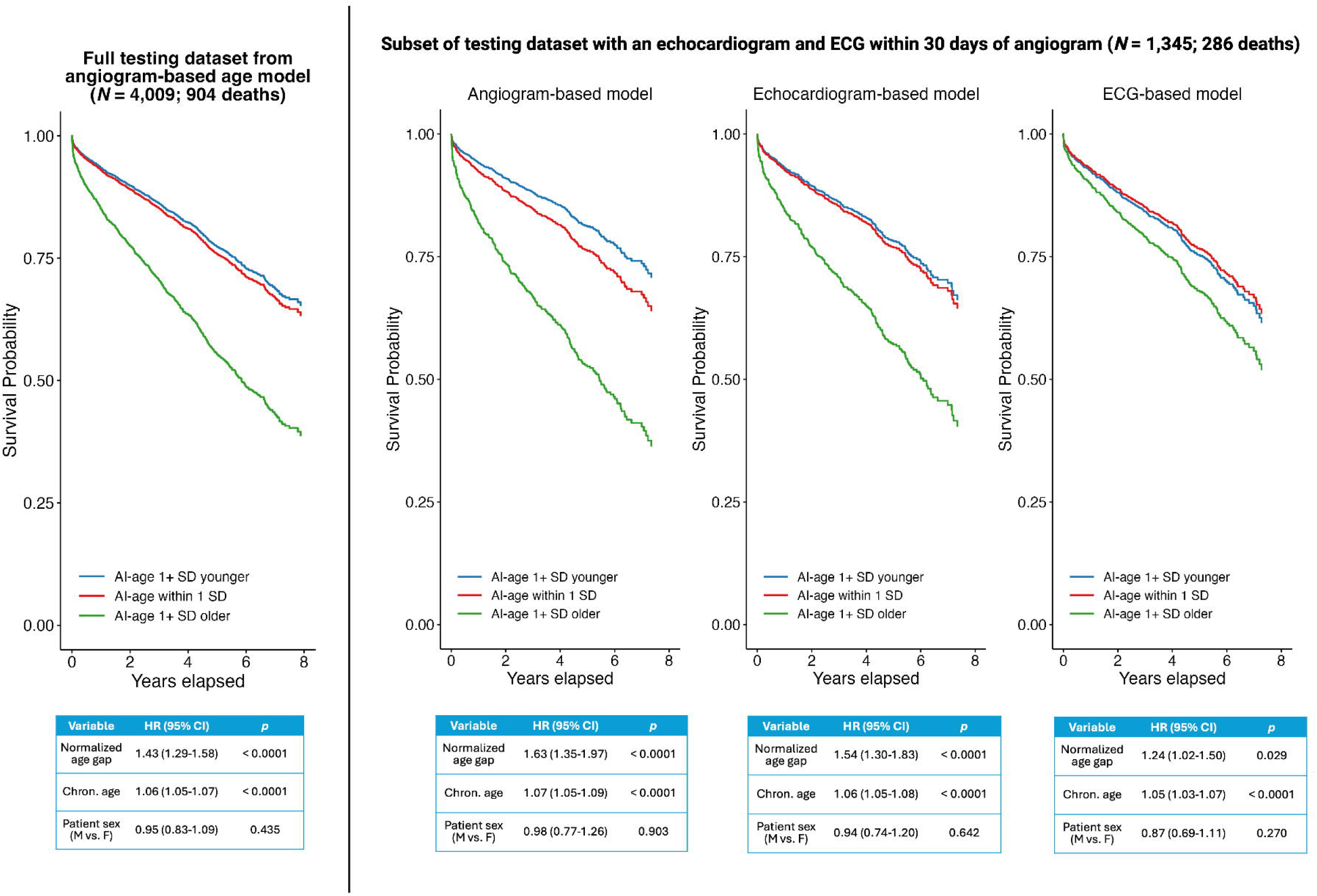
Survival outcomes for the full testing dataset from the angiogram-based model (left) as well as the subset with all three modalities (right). In the survival curves, patients are grouped according to whether their AI-estimated age was within 1 standard deviation (SD) of their chronological age, or greater than 1 SD older or younger than their chronological age. Below the curves, the hazard ratios are from Cox proportional hazards models in which the normalized value of the discrepancy between AI-estimated age and chronological age (the normalized “age gap”) is instead treated as a continuous regressor. All models included patient age and sex (with female as the reference level) as additional regressors. chron. = chronological; CI = confidence interval

When the normalized age gap was averaged across the three modalities to create a multi-modal cardiac aging model (Figure 6), the composite model HR was 2.24 (1.71-2.92, *p* < 0.0001). This pattern is also reflected in the survival curves, as the survival curve is steepest for the group of patients for whom at least two of the three modalities estimated the patient’s cardiac age to be at least 1 SD older than the patient’s chronological age.

**Figure 6.**
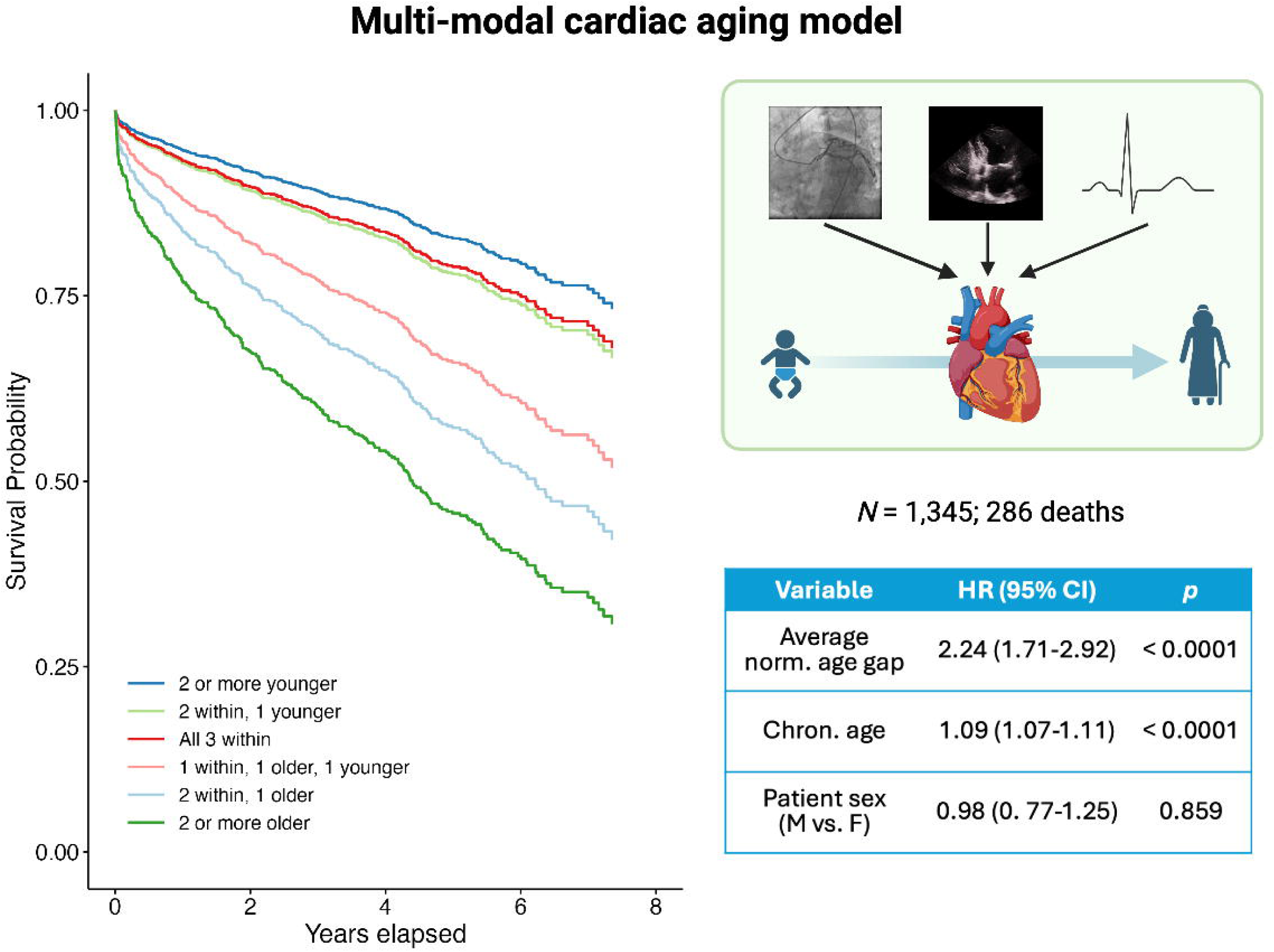
Patient survival outcomes using a multi-modal model of cardiac aging that combined results across angiogram-, echocardiogram-, and ECG-based aging models. For the plots, patients were grouped according to whether all three modalities had an AI-estimated age within 1 SD of the patient’s chronological age, or whether two out of the three had age within 1 SD but the third modality estimated the patient to be at least 1 SD older (or younger) than their chronological age, and so on. To the right of the curves, the hazard ratios are from Cox proportional hazards models in which the average normalized value of the discrepancy between AI-estimated age and chronological age (the average normalized “age gap” across the three modalities) is instead treated as a continuous regressor. Again, patient age and sex (female treated as the refence level) were included as additional regressors for both the model based on patient groupings according to SDs (i.e., shown in the survival curves) as well as the model treating the average normalized age gap as a continuous regressor (shown in the table). The icons for the ECG, heart, infant, and older adult were taken from BioRender.com norm. = normalized; chron. = chronological; CI = confidence interval

We also repeated the multi-modal analysis for female and male patients separately, with results displayed in Table 2. As shown in the table, the HR for the average normalized age gap was considerably higher for female patients (HR = 2.81, 95% CI: 1.82-4.35, *p* < 0.0001) compared to male patients (HR = 1.94, 95% CI 1.38-2.74, *p* = 0.0001). However, this effect, which may have been driven in part by differences in the prevalence of certain comorbidities between the sexes, should be interpreted with caution given the imbalance in the number of patients across the two groups.

**Table 2.** Survival outcomes based on multi-modal models of cardiac aging evaluated separately for female and male patients.

| **Variable** | **Female Patients**<br>**(N = 478; 114 deaths)** |  | **Male Patients**<br>**(N = 867; 172 deaths)** |  |
| --- | --- | --- | --- | --- |
|  | **HR (95% CI)** | ***p*** | **HR (95% CI)** | ***p*** |
| Average norm.<br>age gap | 2.81 (1.82-4.35) | < 0.0001 | 1.94 (1.38-2.74) | 0.0001 |
| Chron. age | 1.09 (1.06-1.13) | < 0.0001 | 1.09 (1.06-1.12) | < 0.0001 |

CI = confidence interval; norm. = normalized; chron. = chronological

In the Supplementary Materials, we also show that the multi-modal aging results held for patients without prior cardiovascular procedures (namely, prostheses, CABG, devices and/or stents), albeit with a slightly lower HR (*N* = 943; HR = 2.06, 95% CI of 1.48-2.89, *p* < 0.0001).

Finally, in addition to examining the average normalized age gap across all three of the angiogram-, echocardiogram-, and ECG-based models, we also examined effects across pairs of modalities. Figure 7 shows the hazard ratios (and 95% confidence interval of the HR) for Cox proportional hazards models evaluating the effect of the normalized age gap for either each modality separately, or the average normalized age gap across pairs of modalities or across all three modalities. These results are provided for all patients as well as separately for female and male patients. Note that for the models with all patients, both chronological age and patient sex were included as additional regressors, whereas for the separate models for female and male patients, only chronological age was included as an additional regressor. As shown in the figure, although the pattern differed between the sexes (likely due to differences in the prevalence of comorbidities), in general the HR increased as more modalities were added.

**Figure 7.**
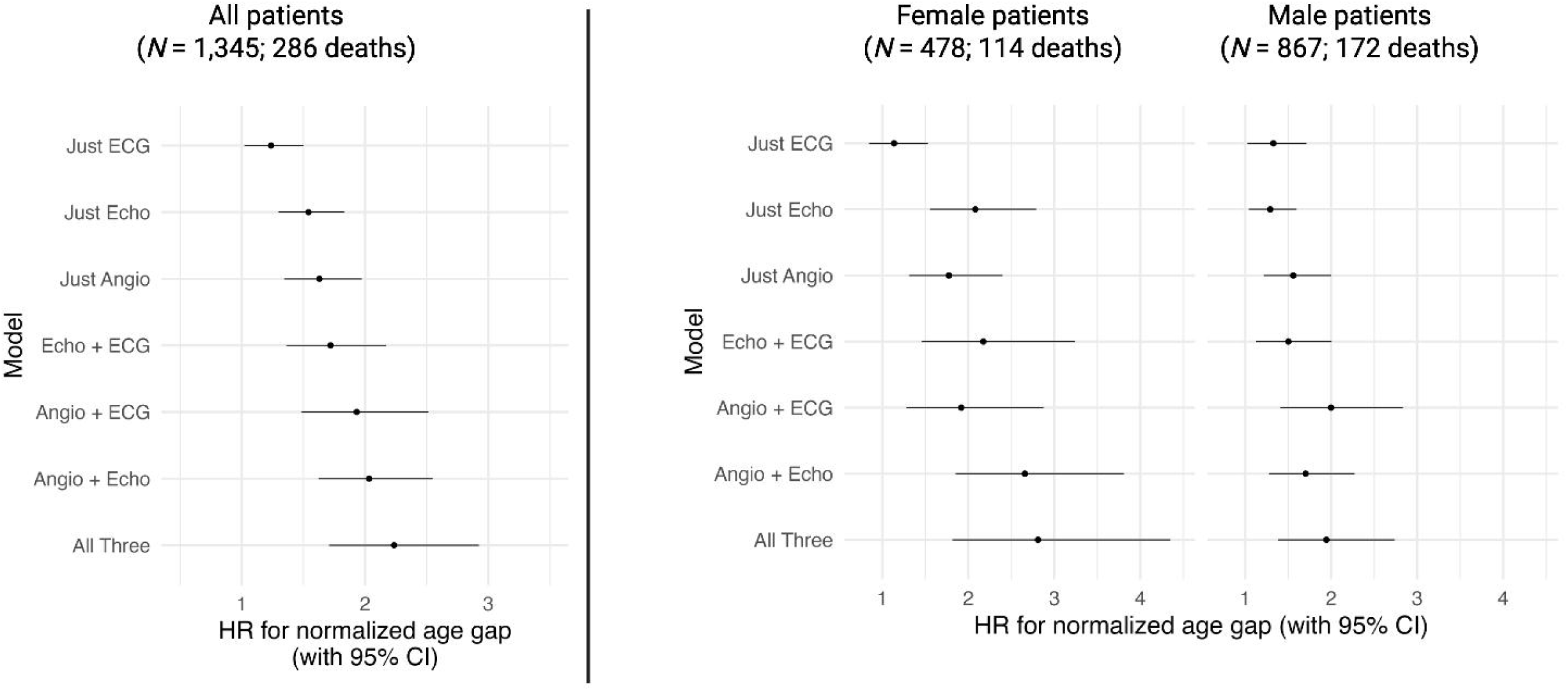
Hazard ratios for the normalized age gap (or the averaged normalized age gap of more than one modality) across combinations of modalities, either for all patients (left) or separately for female and male patients (right).

## Discussion

In this study we found that AI estimates of cardiac age using multimodal inputs including coronary angiograms, echocardiograms, and ECGs were more powerful than estimates from individual modalities alone. This likely results from the added ability of the multimodal model to account for the impact of aging on vascular changes (angiography), structural changes (echocardiography), and electrophysiologic processes (ECG). Physiologic aging (as opposed to chronologic aging) reflects an organism response to environmental stressors over time and the robustness of its repair mechanisms [21]. Understanding physiologic age may permit more accurate prediction (and potentially prevention) of age-related disease, assessment of frailty and risk or benefit of invasive procedures, and evaluation of life-extending therapies. Current assessments of biological aging such as measuring DNA methylation, telomerase activity, and circulating senescent cells involve expensive laboratory assessment of biologic samples not routinely available in clinical practice [21]. The application of AI to data generated as part of routine care facilitates assessment of aging in practice and for population studies. As we gain a richer understanding of inter-individual differences in physiological aging, interventions targeting aging will continue to emerge. Reliable tools to quantify the progression of aging as well as response to treatment will be essential. We found that a multimodal model using inputs that capture different biological aspects of humans enhances biological age determination.

In the present work, we first developed deep learning models to estimate cardiac age and from routine coronary angiograms, which was a novel strategy. The models performed well – with metrics on par with prior work using ECG and echocardiographic data [1, 2, 4, 9] – confirming that vascular imaging contains biological signals relevant to aging. But predicting chronological age is not the objective; it’s merely a training target. The true test of these models lies in whether their predictions relate to patient outcomes. In that sense, the clinical value of these models lies not in how closely they track the calendar, but rather in how they deviate from it, with the mismatch reflecting physiological aging.

The finding that angiographic images predict cardiovascular events and mortality when scrutinized by non-linear neural networks capable of recognizing multiple, likely subtle changes in vessels despite the heterogeneity of views and individual anatomy, comports with our understanding of biology. Stressors that accelerate cardiovascular biological aging include inflammatory conditions, hypertension, diabetes mellitus, tobacco use and other conditions.

These result in identifiable vascular changes including diffuse narrowing, tortuosity, calcification, ulceration, and intimal hyperplasia, as well as regional wall motion abnormalities, all of which may be used by the network to characterize aging [22–24]. While some of these changes (for example, regional wall motion abnormalities) may be detected by other modalities, others (such as vascular stenosis without occlusion or calcifications) are generally not well seen by echocardiography or electrocardiography, resulting in the additive impact of the different modalities [25].

When comparing AI-estimated age across modalities (using the newly developed angiogram-based models and previously developed echocardiogram- and ECG-based models), we observed only moderate agreement. We speculate that this lack of concordance is not a flaw: it confirms that each modality captures a different aspect of the heart’s biology, with electrical, structural, and vascular systems aging at different rates, as noted above. That diversity is precisely what makes a multi-modal model more powerful. When we examined the impact of the discrepancy between patient age and model-estimated age on patient survival outcomes, considering the three modalities in tandem accounted for patient survival with a larger hazard ratio compared to considering each modality in isolation. This finding suggests that by leveraging the complementarity of the different modalities and considering what each technique does and does not capture, future research could potentially use these models to triangulate sources of cardiac aging [10, 11]. Indeed, the pathophysiology of electrophysiology with aging includes degradation of gap junctions and connexons, interstitial fibrosis, and alterations in channel expression that are distinct and in some cases independent from vascular changes [26], whereas other mechanisms (inflammation) may have more in common [27]. Similarly, echocardiography may be affected by tissue changes altering ultrasound backscatter [28]; regional wall motion abnormalities consequent to ischemia, infarction, fibrosis, or other pathophysiologies; calcification of the mitral annulus or aortic valve; and other structural changes. Furthermore, if aging is modifiable, the combined discrepancy across modalities may become the metric we use to measure the specific effects of various interventions [21].

Our findings are best interpreted in light of their limitations. Namely, there is considerable selection bias in the current patient cohort, as all patients had cardiac conditions that necessitated coronary angiography. Therefore, although the patient cohorts upon which the current models were trained and evaluated have a broad age range, they also have a particularly high prevalence of cardiac disease. This selection bias may explain why performance of the ECG-based age estimation model was inferior compared to that of previous studies [29], as the current cohort had very high prevalence rates for a number of cardiac comorbidities that may have impacted AI-ECG performance [1]. Related to this, because the patient cohorts are representative of who receives cardiac testing in our practice, as opposed to the general population, there are likely differences between these cohorts and the general population including differences in race, ethnicity, and sex. To address this limitation, future work should focus on model validation with diverse patient cohorts.

## Conclusion

Measures of biological age may provide insights into the risk of chronic disease, cardiovascular morbidity, and mortality, and guide preventive therapies. We developed deep learning algorithms that showed strong performance in age estimation using routine coronary angiograms as input, and found that a multimodal approach to estimating biological age utilizing ECG, echocardiography, and angiography surpasses any single model in predicting long-term outcomes.

## Code availability

Any custom code or software used to implement the deep learning models detailed in this paper will be made available upon request.

## Data availability

The data are not publicly available because they are electronic health records. Sharing these data externally without additional consent might compromise patient privacy and would violate the Institutional Review Board approval of the study. If other investigators are interested in performing additional analyses, requests can be made to the corresponding author, Z.I.A., and analyses could be performed in collaboration with the Mayo Clinic.

## References

1. Attia, Z.I., et al., Age and Sex Estimation Using Artificial Intelligence From Standard 12-Lead ECGs. Circ Arrhythm Electrophysiol, 2019. 12(9): p. e007284.

2. Ghorbani, A., et al., Deep learning interpretation of echocardiograms. NPJ Digit Med, 2020. 3: p. 10.

3. Duffy, G., et al., Confounders mediate AI prediction of demographics in medical imaging. NPJ Digit Med, 2022. 5(1): p. 188.

4. Anisuzzaman, D., et al., Leveraging comprehensive echo data to power AI models for handheld cardiac ultrasound. Mayo Clinic Proceedings: Digital Health, 2025. 3(1): p. 100194.

5. Kalyakulina, A., et al., eXplainable Artificial Intelligence (XAI) in aging clock models. arXiv, 2023.

6. Avram, R., et al., Automated Assessment of Cardiac Systolic Function From Coronary Angiograms With Video-Based Artificial Intelligence Algorithms. JAMA Cardiol, 2023. 8(6): p. 586–594.

7. Rostami, B., et al., Deep learning to estimate left ventricular ejection fraction from routine coronary angiographic images. JACC: Advances, 2023. 2(9).

8. Le Goallec, A., et al., Predicting arterial age using carotid ultrasound images, pulse wave analysis records, cardiovascular biomarkers, and deep learning. medRxiv, 2021.

9. Anisuzzaman, D., et al., A View-agnostic deep learning framework for comprehensive analysis of 2D echocardiography. medRxiv, 2025.

10. Le Goallec, A., et al., Dissecting heart age using cardiac magnetic resonance videos, electrocardiograms, biobanks, and deep learning. medRxiv, 2021.

11. Siontis, G., et al., Multi-modality deep learning prediction of heart age: Insights from UK-Biobank. European Heart Journal, 2024. 45(October 2024): p. ehae666.3442.

12. He, K., et al. Deep residual learning for image recognition. in Proceedings of the IEEE conference on computer vision and pattern recognition. 2016.

13. Contributors, P. ReduceLROnPlateau. [cited 2025 July 15]; Available from: https://docs.pytorch.org/docs/stable/generated/torch.optim.lr_scheduler.ReduceLROnPlateau. html.

14. Kingma, D.P. and J. Ba, Adam: A method for stochastic optimization. arXiv preprint arXiv, 2014.1412.6980.

15. Paszke, A., et al., Pytorch: An imperative style, high-performance deep learning library. Advances in Neural Information Processing Systems, 2019. 32.

16. Fawcett, T., An introduction to ROC analysis. Pattern recognition letters, 2006. 27(8): p. 861–874.

17. Naser, J.A., et al., Artificial intelligence-based classification of echocardiographic views. European Heart Journal - Digital Health, 2024. 5(3): p. 260–269.

18. Therneau, T.M., A Package for Survival Analysis in R. 2024.

19. Kassambara, A., M. Kosinski, and P. Biecek, survminer: Drawing Survival Curves using ’ggplot2’. 2021.

20. Denz, R., R. Klaaßen-Mielke, and N. Timmesfeld, A comparison of different methods to adjust survival curves for confounders. Statistics in Medicine, 2023. 42(10): p. 1461–1479.

21. Lopez-Jimenez, F., et al., Assessing Biological Age: The Potential of ECG Evaluation Using Artificial Intelligence: JACC Family Series. JACC Clin Electrophysiol, 2024. 10(4): p. 775–789.

22. Guzik, T.J. and R.M. Touyz, Oxidative stress, inflammation, and vascular aging in hypertension. Hypertension, 2017. 70(4): p. 660–667.

23. Tesauro, M., et al., Arterial ageing: from endothelial dysfunction to vascular calcification. J Intern Med, 2017. 281(5): p. 471–482.

24. Sutton, N.R., et al., Molecular Mechanisms of Vascular Health: Insights From Vascular Aging and Calcification. Arterioscler Thromb Vasc Biol, 2023. 43(1): p. 15–29.

25. Saxena, A., E.Y.K. Ng, and S.T. Lim, Imaging modalities to diagnose carotid artery stenosis: progress and prospect. Biomed Eng Online, 2019. 18(1): p. 66.

26. Horn, M.A., Cardiac Physiology of Aging: Extracellular Considerations. Compr Physiol, 2015. 5(3): p. 1069–121.

27. Wang, X., et al., The immune system in cardiovascular diseases: from basic mechanisms to therapeutic implications. Signal Transduct Target Ther, 2025. 10(1): p. 166.

28. Masuyama, T., et al., Ultrasonic tissue characterization with a real time integrated backscatter imaging system in normal and aging human hearts. J Am Coll Cardiol, 1989. 14(7): p. 1702–8.

29. Ladejobi, A.O., et al., The 12-lead electrocardiogram as a biomarker of biological age. European Heart Journal-Digital Health, 2021. 2(3): p. 379–389.

