## Supplementary Material for "Seeing the Aging Heart: Multimodal AI Quantifies Cardiac Biological Aging from Angiography, Echocardiography, and ECG"

***Table of Contents***

Supplementary Methods 2

Identification of patients with prior cardiovascular procedures 2

Model hyperparameters 3

Supplementary Results 3

Sensitivity analyses based on prior cardiovascular procedures 3

Supplementary Table 1. Percentage of patients with prior cardiovascular procedures in each of the patient cohorts 4

Supplementary Figure 1. Survival curves for individual modalities for patients without cardiovascular procedures 4

Supplementary Figure 2. Survival curves for a multi-modal cardiac aging model for patients without cardiovascular procedures 5

References 6

### Supplementary Methods

#### Identification of patients with prior cardiovascular procedures

Prosthesis and coronary artery bypass grafting (CABG) information were obtained from structured echocardiography reports. Each prosthetic device was classified into one of three categories: transcatheter (including aortic, mitral, tricuspid, and pulmonary valve replacements, as well as edge-to-edge repair devices), mechanical (such as bileaflet or disc prosthetic valves), or surgical tissue (including bioprostheses, homografts, conduits, annuloplasty rings, bands, or patch enlargements). For each angiographic study, all prior echocardiograms were reviewed, and any documented prosthetic procedures or CABG were aggregated and recorded as prior exposure. Both the devices and procedures, along with their assigned categories, were listed to ensure that each angiographic study was annotated with the complete history of prosthetic interventions and CABG available from preceding echocardiography.

Device and stent information was obtained by linking each angiographic study to its associated hardware records. Each device was classified into one of five categories based on the recorded hardware type: percutaneous coronary interventions (PCI; coronary stents and related procedures including percutaneous transluminal coronary angioplasty, atherectomy, thrombectomy, and laser), rhythm (implantable cardioverter–defibrillators, pacemakers, and associated leads or loop recorders), valve/structural (transcatheter or structural cardiac implants such as valves and left atrial appendage or patent foramen ovale closure devices), vascular (non-coronary vascular grafts or other vascular implants), and other/legacy (devices flagged as legacy or uncategorized in the registry). For each angiographic study, all device entries available for that patient up to and including the study date were reviewed, and both the individual devices and their assigned categories were summarized as prior device exposure.

**Model hyperparameters**

Each model was trained for 50 epochs, and the value of 1e-5 was used for the initial learning rate, which was then decayed during training using the *ReduceLROnPlateau* strategy [1]. A value of 0.001 was selected for weight decay, and the mini-batch size was set to 64. The Adam algorithm [2] was utilized for optimization, and the loss functions was Root Mean Square Error (RMSE). All experiments and models were implemented in the Python programming language (version 3.10.4) and on the PyTorch (version 1.12.1+cu116) platform [3] using two NVIDIA GA102GL [A10] GPUs.

### Supplementary Results

**Sensitivity analyses based on prior cardiovascular procedures**

Of the 4,009 patients in the testing cohort for the angiogram-based model as well as the subset of 1,345 patients with all three modalities, we performed sensitivity analyses by restricting cohorts to patients who did not have prior cardiovascular procedures (namely, prostheses, CABG, devices and/or stents; percentages of patient cohorts reported in Supplementary Table 1). This resulted in 2,856 patients for the testing cohort for the angiogram-based model and 943 patients with all three modalities.

As shown in Supplementary Figure 1 (for each modality in isolation) and Supplementary Figure 2 (using a multi-modal approach), survival outcomes were comparable to those reported with the full sample, albeit with lower HR values across the board (note that for ECG on its own, the HR for the normalized age gap became insignificant in this smaller cohort).

**Supplementary Table 1.** Percentage of patients with prior cardiovascular procedures in each of the patient cohorts.

| | **Angiogram Model Development** | | | **Subset of testing dataset with echo and ECG within 30 days of angiogram** |
|---|---|---|---|---|
| | **Training** | **Validation** | **Testing** | |
| Prior cardiovascular procedures (%) | | | | |
| Prostheses | 6.14 | 5.86 | 6.30 | 6.84 |
| CABG | 2.55 | 2.57 | 2.71 | 2.53 |
| Devices and/or stents | 21.62 | 21.74 | 21.77 | 22.90 |

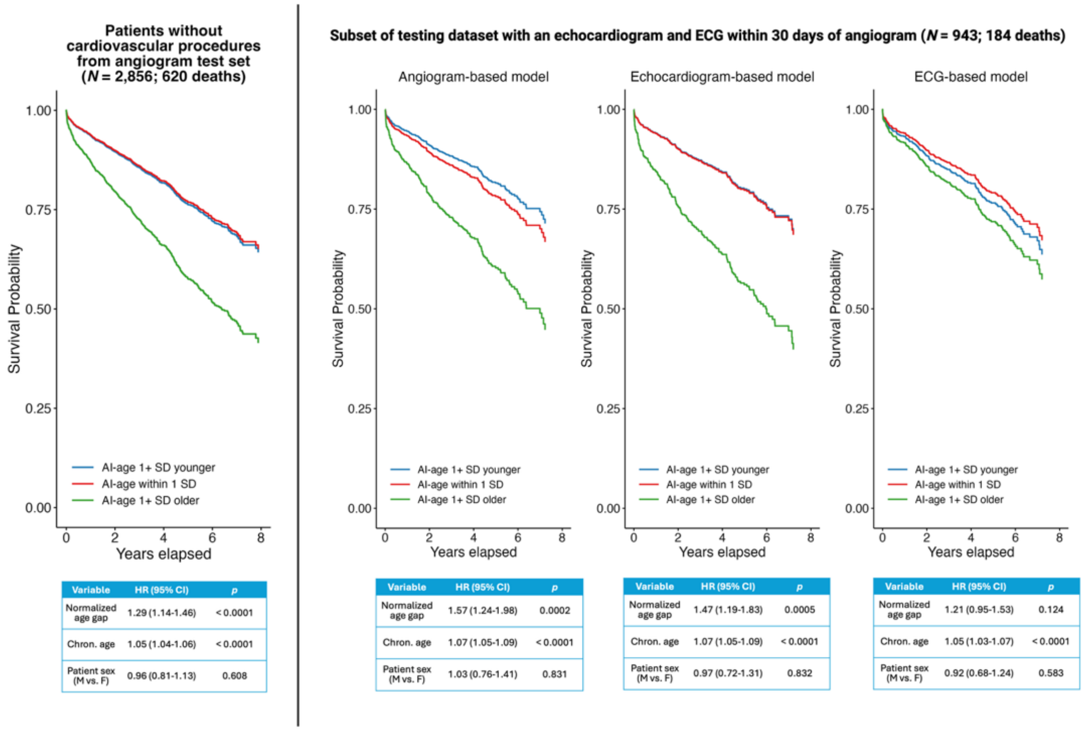


**Supplementary Figure 1.** Survival outcomes for patients without cardiovascular procedures, both from the testing dataset from the angiogram-based model (left) as well as the subset with all three modalities (right). In the survival curves, patients are grouped according to whether their AI-estimated age was within 1 standard deviation (SD) of their chronological age, or greater than 1 SD older or younger than their chronological age. Below the curves, the hazard ratios are from Cox proportional hazards models in which the normalized value of the discrepancy between AI-estimated age and chronological age (the normalized "age gap") is instead treated as a continuous regressor. All models included patient age and sex (with female as the reference level) as additional regressors.
chron. = chronological; CI = confidence interval

Multi-modal cardiac aging model

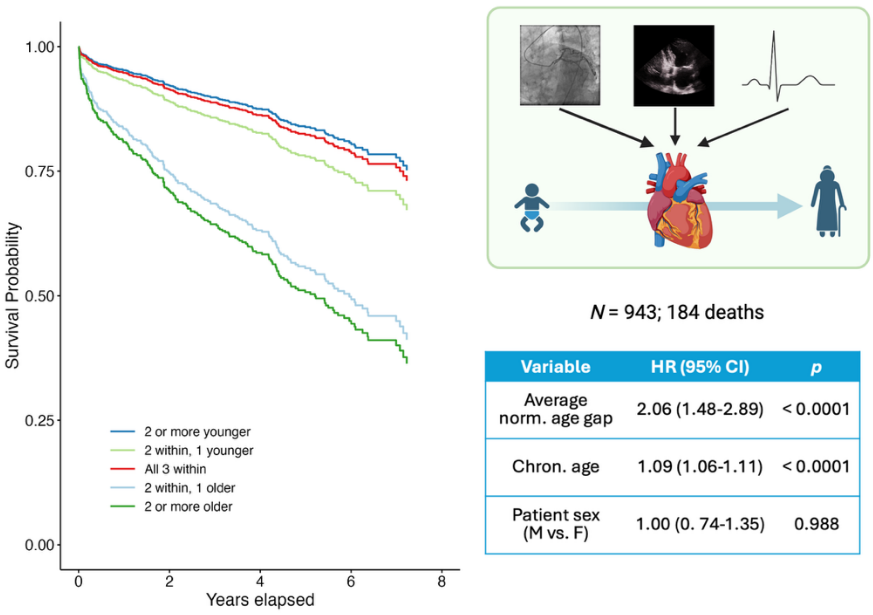


| Variable | HR (95% CI) | *p* |
|---|---|---|
| Average norm. age gap | 2.06 (1.48-2.89) | < 0.0001 |
| Chron. age | 1.09 (1.06-1.11) | < 0.0001 |
| Patient sex (M vs. F) | 1.00 (0. 74-1.35) | 0.988 |

**Supplementary Figure 2.** Patient survival outcomes using a multi-modal model of cardiac aging that combined results across angiogram-, echocardiogram-, and ECG-based aging models for the subset of patients without cardiovascular procedures. For the plots, patients were grouped according to whether all three modalities had an AI-estimated age within 1 SD of the patient's chronological age, or whether two out of the three estimated age within 1 SD but the third modality estimated the patient to be at least 1 SD older (or younger) than their chronological age, and so on. Note that for the purposes of visualization, the group with "1 within, 1 younger, and 1 older" was removed because there were only 9 patients in the group. To the right of the curves, the hazard ratios are from Cox proportional hazards models in which the average normalized value of the discrepancy between AI-estimated age and chronological age (the average normalized "age gap" across the three modalities) is instead treated as a continuous regressor. Again, patient age and sex (female treated as the refence level) were included as additional regressors for both the model based on patient groupings according to SDs (i.e., shown in the survival curves) as well as the model treating the average normalized age gap as a continuous regressor (shown in the table). The icons for the ECG, heart, infant, and older adult were taken from BioRender.com
norm. = normalized; chron. = chronological; CI = confidence interval

### References

1. Contributors, P. *ReduceLROnPlateau*. [cited 2025 July 15]; Available from: https://docs.pytorch.org/docs/stable/generated/torch.optim.lr_scheduler.ReduceLROnPlateau.html.
2. Kingma, D.P. and J. Ba, *Adam: A method for stochastic optimization.* arXiv preprint arXiv, 2014. **1412.6980**.
3. Paszke, A., et al., *Pytorch: An imperative style, high-performance deep learning library*. Advances in Neural Information Processing Systems, 2019. **32**.
